# High-intensity progressive resistance training to prevent falls in community-dwelling adults aged 80 years and older with reduced muscle strength: protocol for the ReFit randomised controlled trial

**DOI:** 10.64898/2026.09.23.26363212

**Authors:** Jonathan Berg, Karina Hammer Tømmerdal, Jeff S. Coombes, Turid Follestad, Øyvind O. Salvesen, Erik Madssen, Geir Selbæk, Håvard Dalen, Gudrun M. W. Bjørnelv, Brooke L. Devlin, Aslak Steinsbekk, Dorthe Stensvold, Maria Fiatarone Singh, Ulrik Wisløff

## Abstract

**Introduction:** Falls are a major cause of injury-related morbidity, mortality, and healthcare utilisation among older adults, with the greatest burden occurring in those aged ≥80 years. Multicomponent exercise programmes that include balance and resistance exercises reduce falls, whereas the effect of resistance training delivered as a standalone fall-prevention intervention remains uncertain. Existing resistance-training trials have generally been small, underpowered for fall outcomes, and have produced inconsistent findings, despite reduced muscle strength being an important modifiable risk factor in the oldest old. The resistance training prescription used in these studies has not always aligned with recommendations for optimal strength adaptations in terms of intensity or progressive overload, which may explain this uncertain efficacy. Therefore, this study aims to determine whether 12 months of high-intensity progressive resistance training reduces fall rates in community-dwelling adults aged ≥80 years with reduced muscle strength.

**Methods and analysis:** This single-centre, randomised controlled trial conducted in Trondheim, Norway, includes 241 community-dwelling adults aged ≥80 years with reduced muscle strength. Participants are randomly assigned (1:1) to either standard care with written physical activity guidance or a twice-weekly high-intensity progressive resistance training intervention for 12 months. The primary outcome is the rate of falls over 12 months. Secondary outcomes include time to first fall, fall-related injuries, muscular function, physical performance, balance, body composition, cardiometabolic health, cognition, and participant-reported outcomes. Cost-effectiveness is assessed from a healthcare perspective over the 12-month intervention period using costs and quality-adjusted life years. Analyses follow the intention-to-treat principle, using negative binomial regression for the primary outcome. The first participant was included on 30 March 2023, and the last participant was included on 29 September 2025. The 12-month intervention period is expected to be completed by September 2026.

**Ethics and dissemination:** The study has received approval from the Regional Committee for Medical and Health Research Ethics in Norway (REK; reference number 462261). All participants provided written informed consent before participating. Findings will be shared via peer-reviewed publications, conference presentations, and communication with stakeholders and the public.

**Trial registration number:** ClinicalTrials.gov (NCT05691166), registered 11 January 2023. Available at: https://clinicaltrials.gov/study/NCT05691166

## INTRODUCTION

The global population is ageing, and the World Health Organisation reports that the number of older adults aged ≥80 years will triple between 2020 and 2050.^1^ Falls among older adults are a major public health challenge, contributing substantially to morbidity, mortality, and healthcare utilisation.^2^ ^3^ The burden of falls increases markedly with advancing age, with the oldest age groups having the highest risk of fall-related mortality.^4^

The increased risk of falls among the oldest old is multifactorial, reflecting an accumulation of chronic diseases, sensory impairments, cognitive decline, and age-related decline in muscle strength.^5–8^ Muscle strength diminishes progressively with age, and this decline is central to the concept of sarcopenia — a clinical condition characterised by low muscle strength and mass that is strongly associated with an increased risk of falls and fall-related fractures.^9^ ^10^As both low muscle strength and falls are common in the oldest old, muscle weakness represents a key modifiable target for preventive measures.^7^ ^8^ ^11^

Exercise effectively reduces falls, with the strongest evidence supporting multicomponent programmes incorporating balance and resistance training (RT).^12–14^ In contrast, the independent effect of resistance training on falls remains uncertain.^13^ Previous trials often used substandard training protocols not concordant with current RT recommendations for optimal strength adaptation,^15^ short intervention periods, or unsupervised home-based exercises, and frequently failed to yield significant gains in muscle strength.^16–18^ Trials that have achieved meaningful muscle strength gains using high-intensity resistance training have shown more promising effects on fall risk, but have generally been underpowered to provide conclusive evidence.^19–21^ Accordingly, current evidence on resistance training as a standalone fall prevention strategy remains limited and of low certainty. There is substantial evidence that resistance training is safe for older adults, including the oldest old, although myths about potential harms remain a common barrier to participation amongst practitioners and consumers.^22^ ^23^

High-intensity progressive resistance training is the approach most consistently shown to produce meaningful gains in muscle strength, and may therefore be particularly important for reducing falls.^15^ This is especially relevant for individuals with reduced muscle strength, in whom muscular weakness may directly contribute to fall risk.^24–26^ Strength gains from resistance training are also linked to improved balance, providing a plausible mechanism by which the intervention may reduce falls.^25^ In addition to improving muscle strength, resistance training may benefit physical function, cardiometabolic health, cognitive function, and mental health, which may further reduce fall risk indirectly.^27–29^

To our knowledge, this is the first randomised controlled trial to investigate the effects of 12 months of high-intensity progressive resistance training on fall outcomes in community-dwelling adults aged ≥80 years with reduced muscle strength, compared with a control group. The study also evaluates secondary health outcomes and cost-effectiveness, and characterises adverse events and potential harms associated with the intervention.

## METHODS AND ANALYSIS

### Patient and public involvement

The development of the Reducing Falls with Progressive Resistance Training trial (ReFit) drew on extensive experience from previous exercise studies involving older adults, including the Generation 100 Study,^30^ a large randomised aerobic exercise trial in older adults, dialogue with older adults and relevant community and older-adults organisations on recruitment strategy, participation in research, exercise delivery, and barriers to long-term engagement.

During the specific planning of ReFit, we conducted interviews with older adults attending fitness centres. Interviewees were informed about the proposed resistance-training trial for adults aged ≥80 years and asked to provide feedback on recruitment materials, delivery mode, supervision, and practical aspects of participation. We recognise that these individuals represented a relatively active and exercise-engaged group, and their perspectives were therefore considered alongside the broader experience gained from engagement with older adults and relevant stakeholders.

During implementation, we have continued to review study procedures and practical experiences with user representatives, including older adults with prior experience of participating in exercise research, and with clinical and community stakeholders, including a physician, a physiotherapist, and a representative from the Norwegian Health Association.

Their feedback informed selected adaptations to study design and delivery, including post-trial exercise provision for control participants after completion of the 12-month primary outcome follow-up, and the option for intervention participants to complete one weekly session without supervision during the second half of the intervention. These adaptations were intended to support retention and longer-term exercise participation.

### Trial design and setting

ReFit is a single-centre, parallel-group randomised controlled superiority trial with a 1:1 allocation ratio, conducted in Trondheim and neighbouring municipalities in Central Norway. The study is coordinated by the Norwegian University of Science and Technology (NTNU) in collaboration with St. Olav’s University Hospital, and the exercise intervention is delivered free of charge to participants at nine community-based fitness centres in Trondheim, operated in collaboration with a commercial fitness centre chain. Sites are selected individually for each participant in the intervention group, based on their place of residence and training preference. The trial protocol is reported in accordance with the updated recommendations of the Standard Protocol Items for Randomised Trials (SPIRIT) statement.^31^

### Eligibility criteria

Table 1 provides the complete eligibility criteria. Community-dwelling adults aged 80 years or older are eligible if they have reduced muscle strength, operationalised according to the 2019 European Working Group on Sarcopenia in Older People (EWGSOP2) concept of probable sarcopenia.^32^ In EWGSOP2, reduced muscle strength constitutes probable sarcopenia and is considered sufficient in clinical practice to trigger initiation of intervention. In ReFit, reduced muscle strength is defined as meeting either of two criteria: low grip strength or impaired five-times chair-stand performance. The chair-stand test is a reasonable proxy for lower-limb strength, although performance also depends on other components of physical function.^33^ Low grip strength is defined as a T-score below -2.0 relative to sex-specific peak grip strength values in a healthy Norwegian adult reference population, corresponding to <41.6 kg in men and < 23.4 kg in women,^34^ while impaired chair-stand performance is defined as taking >15 seconds to complete five chair rises without using the arms. The grip strength criterion follows the EWGSOP2 recommendation to use normative reference values where available, whereas the chair-stand criterion uses the EWGSOP2-specified cut-off. These criteria are selected to identify older adults with probable sarcopenia while maintaining recruitment feasibility in a community-based trial. Exclusion criteria are listed in Table 1.

**Table 1.** Study eligibility criteria.

| Inclusion criteria | Exclusion criteria |
| --- | --- |
| Age ≥ 80 years | Pre-existing diagnosis of dementia |
| Reduced muscle strength<sup>†</sup> (≥1 of the following):<br>- Five-times sit-to-stand $>$15.0 s<br>- Grip strength $<$41.6 kg (men)<br>- Grip strength $<$23.4 kg (women) | Moderate or severe cognitive impairment (score $<$18 on the Mini-Mental State Examination)<sup>52</sup> |
| Community-dwelling, including independent senior housing | Living in institutional care |
| Ambulatory without supervision or physical assistance from another person. Assistive devices, such as canes, crutches, or walkers, are allowed. | Non-ambulatory or requiring a person or a wheelchair to assist when walking |
| Able to see and hear sufficiently to undertake assessments and partake in the planned exercise training. | Degenerative neurological and neuromuscular disease/disorder significantly influencing gait and mobility (e.g. amyotrophic lateral sclerosis and Parkinson's disease) |
|  | Amputation (other than toes/fingers). |
|  | Contraindications for resistance training* |
|  | Unstable fracture |
|  | Inability to comply with study requirements |
|  | Currently undertaking progressive resistance training |

\* Examples of contraindications include: unstable coronary heart disease, decompensated heart failure, uncontrolled arrhythmias, severe pulmonary hypertension, severe and symptomatic aortic stenosis, acute myocarditis, endocarditis, pericarditis, uncontrolled hypertension ( $>$ 200/110 mmHg), aortic dissection, Marfan syndrome, diabetic retinopathy, and rapidly progressive terminal illness.<sup>53</sup>
<sup>†</sup>Reduced muscle strength defined in line with the EWGSOP2 concept of probable sarcopenia. The grip-strength threshold is based on sex-specific normative reference values representing peak strength in a healthy Norwegian adult population; the chair-stand threshold is the EWGSOP2-specified cut-off.

### Intervention and comparator

#### Intervention

Participants randomised to the intervention group complete two sessions per week of whole-body high-intensity progressive resistance training for 12 months. The programme comprises five machine-based exercises targeting major muscle groups of the lower and upper body, with particular emphasis on muscles relevant to falls prevention (Figure 2).^35^ Participants perform 24 repetitions per exercise (e.g., 3 sets of 8 or 6 sets of 4 repetitions), with approximately 1-2 minutes of rest between sets, adjusted according to individual recovery.

Training intensity is individually prescribed relative to one-repetition maximum (1RM) for each exercise. Following familiarisation and initial 1RM testing, training loads progressively increase from 50% to 80% of 1RM.^36^ From session 8 onwards, participants train at ≥80% of their most recently measured 1RM. To maintain progressive overload between 1RM reassessment, the load is increased by approximately 1% per session, guided by rating of perceived exertion within the target range of 15-18 on the Borg 6-20 scale. This progression approach is adapted from a previous high-intensity resistance training protocol in older adults using small session-by-session load increments guided by ratings of perceived exertion.^36^ Fractional weight plates are added to the weight stack of the resistance training machines to permit these small increments. The 1RM is reassessed every 12^th^ session to recalibrate the prescribed training load as strength improves, thereby maintaining a progressive high-intensity training stimulus throughout the intervention. Reassessment sessions form part of the intervention schedule and generally replace the usual training prescription for that session. The protocol is designed to optimise improvements in muscular strength.^37^

Training sessions are delivered at the participating community-based fitness centres. For the first six months, instructors supervise all training sessions in groups of up to five participants. Instructors are physiotherapists, exercise physiologists, qualified personal trainers, and individuals holding at least a bachelor’s degree in a health-or exercise-related discipline. All instructors receive study-specific training in the intervention protocol, including exercise selection, progression, safety procedures, supervision, and documentation. After six months, participants continue to complete two training sessions per week. One session may be completed without supervision, whereas at least one session per week must remain supervised. Participants document any unsupervised sessions in a training log, while supervised sessions are documented by the instructor. Standardised training logs record attendance, exercises, training intensity, volume, supervision status and perceived exertion.

Participants in the intervention group are neither encouraged nor discouraged from performing additional exercise beyond the intervention. The intervention may be modified or discontinued in response to adverse events, new or worsening medical conditions, participant request, clinical judgement or medical advice. Where possible, modifications are preferred over discontinuation and may include adjustments to range of motion, exercise substitution or reductions in training intensity. Participants who discontinue the intervention are encouraged to continue outcome assessments. In cases of repeated non-attendance, study staff contact participants to explore reasons for absence and facilitate continued participation where possible. If prolonged absence occurs, training loads may be reassessed and adjusted.

#### Comparator

Participants randomised to the comparator (control) group receive a booklet containing Norway’s current recommendations for physical activity for older adults. These recommendations include ≥150 minutes of moderate-intensity or ≥75 minutes of vigorous-intensity physical activity per week, together with balance and muscle-strengthening exercises 2–3 times per week. The booklet also includes instructions for seven exercises intended to reduce fall risk; these exercises are summarised in Supplementary Table 2. The comparator reflects standard care, in which structured, supervised exercise programmes are typically not provided to this population, thereby allowing evaluation of the intervention under real-world conditions. As in the intervention group, control participants are neither encouraged nor discouraged from engaging in exercise or physical activity beyond the recommendations in the booklet.

We assess exercise behaviour outside the intervention sessions by self-report every three months using a structured questionnaire administered through the eForsk system. Depending on participant preference, participants complete the questionnaire online or study staff administer it by telephone. The definition of overall exercise behaviour (for use in frailty outcome and other analyses) includes both study-related and non-study-related exercise.

### Participant recruitment, screening, and consent

Potential participants aged ≥80 years residing in the trial catchment area are identified from the National Population Register. Between January 2023 and August 2025, invitation letters containing a study information booklet and response sheet are sent to potential participants. The response sheet allows recipients to indicate their interest in participating, and all recipients, regardless of their interest, are asked to return it.

Interested individuals are contacted by telephone and undergo an initial phone screening for demographic information, self-reported medical history, and current exercise participation, in accordance with the trial’s eligibility criteria (Table 1). If interested and eligible after the initial phone screening, individuals are invited and scheduled for an in-person screening visit.

At the in-person screening visit, trained study staff obtain written informed consent before assessing grip strength, chair-stand performance, and the Mini-Mental State Examination (MMSE) to determine whether participants meet the muscle-strength and cognitive eligibility criteria specified in Table 1. All attendees also complete the full Short Physical Performance Battery (SPPB) and a body-composition assessment to characterise the broader screening population. Individuals who meet the eligibility criteria are then scheduled for a separate assessment day to complete the remaining baseline tests.

### Trial status

Recruitment for the trial was completed on 29 September 2025, with 241 participants randomised. The 12-month intervention and primary outcome follow-up are expected to be completed in September 2026. The extended 24- and 36-month follow-ups are expected to be completed in September 2028.

### Randomisation and blinding

After completing baseline examinations, participants are randomised 1:1 using the eForsk randomisation system (described in further detail under Data collection methods and data management). A computer-generated permuted-block randomisation sequence generated within eForsk is used, stratified by sex, with varying block sizes. The randomisation sequence and upcoming treatment allocations are inaccessible to investigators and study staff. After completing baseline assessments and confirming eligibility, study staff initiate randomisation in eForsk. Treatment allocation remains concealed until the randomisation request is completed, after which the assigned group is displayed on screen and communicated verbally to the participant by study staff.

Participants and exercise instructors cannot be blinded to group allocation due to the exercise intervention. Baseline assessments are completed before randomisation. At follow-up, efforts are made to preserve assessor blinding by assigning assessments, where feasible, to study staff who are unaware of the participant’s allocation and not involved in delivering the intervention to that participant. However, complete assessor blinding cannot be guaranteed because some study staff have roles in both outcome assessment and intervention delivery.

### Outcomes

Assessments are conducted at baseline, 6, 12, 24 and 36 months, with all physical assessments performed at the same time of day for each participant across all time points. Because control participants are offered a fitness-centre membership after the 12-month primary outcome follow-up (see Ancillary and post-trial care), between-group comparisons during the 24- and 36-month extended follow-up are exploratory. Figure 1 provides an overview of study outcomes, including the timing of each assessment.

**Figure 1.**
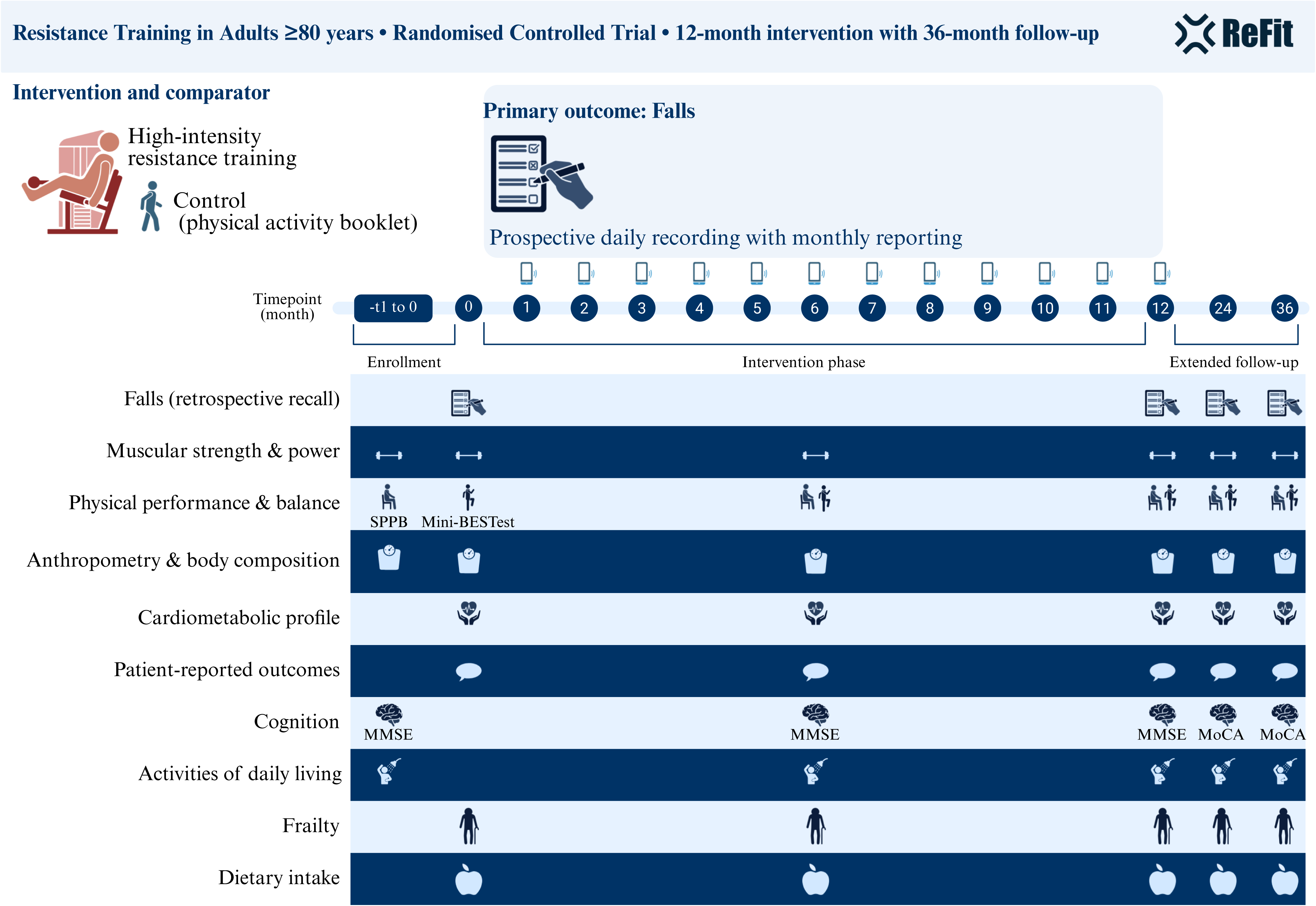
Overview of the ReFit trial design, timeline, and outcome assessment schedule. Community-dwelling adults aged ≥80 years with reduced muscle strength are randomised to high-intensity progressive resistance training or standard care. The primary outcome, rate of falls, is recorded prospectively using daily calendars with monthly reporting throughout the 12-month intervention. Clinical assessments are conducted at baseline, 6 and 12 months, and during extended follow-up at 24 and 36 months. Some assessments were introduced after trial commencement and are not available for all participants at all time points. MMSE, Mini-Mental State Examination; MoCA, Montreal Cognitive Assessment; SPPB, Short Physical Performance Battery.

**Figure 2.**
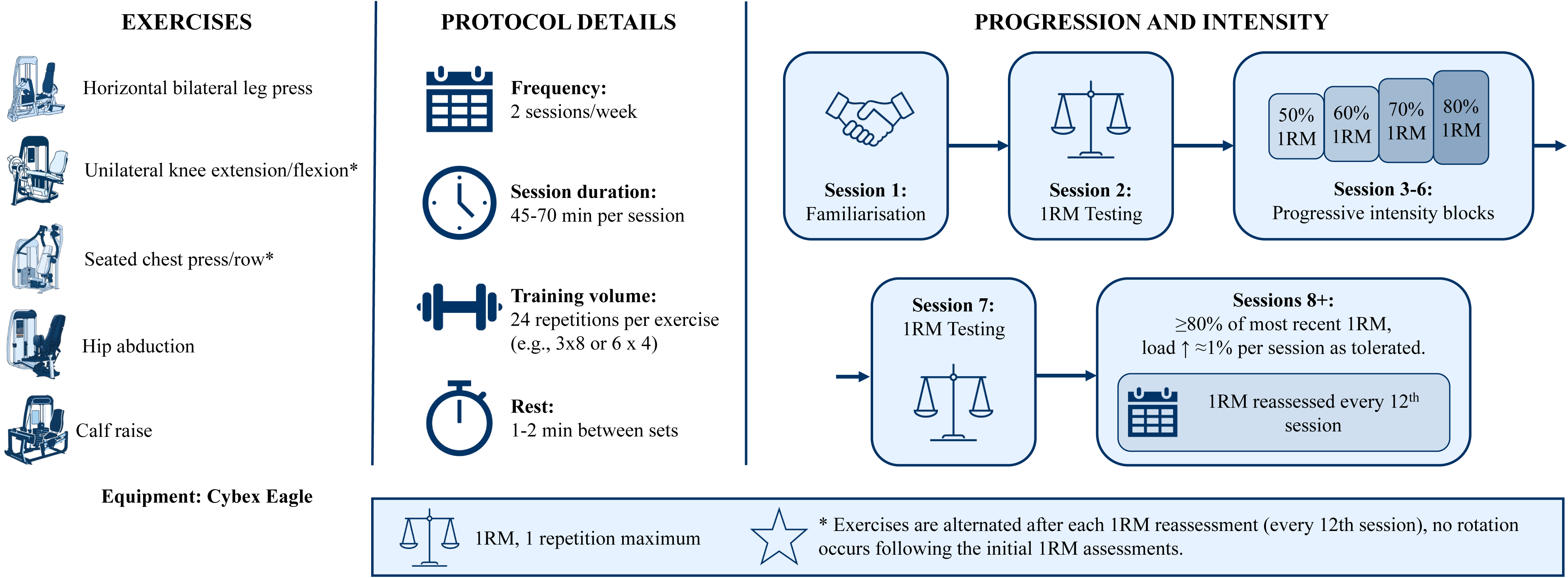
Resistance training protocol: exercises, prescription, and progression. The intervention comprises twice-weekly high-intensity progressive resistance training on resistance machines (Cybex Eagle), targeting major lower- and upper-body muscle groups. Following familiarisation and one-repetition-maximum (1RM) testing, load is progressed from 50–80% 1RM to ≥80% of the most recent 1RM, with 1RM reassessed every 12th session. Asterisked exercises are alternated after each 1RM reassessment. 1RM, one-repetition maximum.

### Primary outcome measure

The primary outcome is the fall rate per person-year during the 12-month intervention period, based on prospectively recorded falls and the time for which fall-outcome data are available. Falls are defined as any unexpected event in which the participant comes to rest on the ground, floor, or lower level, according to the Prevention of Falls Network Europe (ProFaNE).^38^ Falls are prospectively monitored using a printed fall calendar provided at baseline and covering the entire 12-month follow-up period. The calendar comprises 12 monthly date-based grids, and participants are instructed to record daily whether a fall has occurred and, when a fall occurs, to note details about the event.

At the end of each month, participants report falls through an online questionnaire administered via eForsk or a telephone-administered questionnaire with the same questions, per participant preference. Study personnel enter telephone responses directly into eForsk. Study personnel contact participants who choose electronic reporting but do not submit a monthly report to obtain fall information. For each reported fall, information is collected on the date, location, activity and consequences, including whether the fall resulted in injury or required medical attention. For participants unable to complete the calendar or the monthly questionnaire themselves, a proxy (e.g., a family member or caregiver) may do so on their behalf.

The monthly reports and fall calendars are considered complementary sources of fall information. Monthly reports recorded in eForsk constitute the primary electronic record, while the calendars provide a contemporaneous prospective record and are returned at the 12-month assessment for review before finalisation of the fall-outcome dataset. Falls reported through either source are considered for inclusion if they meet the study definition of a fall.

When both sources appear to describe the same event, we use information on the date and circumstances of the fall to identify potential duplicates, and we count the event only once. If the sources differ only in the reported date and clearly refer to the same event, we use the calendar date where available because it represents the contemporaneous record. If a monthly report cannot be obtained, calendar information may be used to ascertain falls for that interval when sufficiently complete; otherwise, the interval is considered to have missing fall data.

### Secondary outcome measures

Secondary outcomes are selected to characterise the intervention’s broader effects on fall-related events, muscular function, physical performance, body composition, cardiometabolic health, cognition, patient-reported outcomes, and healthcare utilisation. Figure 1 summarises the timing of all assessments, and Supplementary Table 1 provides detailed measurement procedures and instruments.

Fall-related secondary outcomes include experiencing one or more falls, time to first fall, the number of falls requiring medical attention, and the number and occurrence of fall-related fractures. Muscle function is assessed using leg-press 1RM, maximal handgrip strength, and submaximal leg-press power. The leg press (M456, Technogym, Italy) used for outcome assessment differs from the leg press (Cybex Eagle) used in the intervention for logistical reasons detailed in Supplementary Table 1. The Technogym M456 is used consistently for outcome assessment in both randomised groups. We assess physical performance and balance using the Short Physical Performance Battery and the Mini Balance Evaluation Systems Test (Mini-BESTest).^39–41^ Anthropometrics and body composition are assessed using standard anthropometric measurements and bioelectrical impedance analysis. We assess physical frailty using Fried’s Frailty Phenotype and operationalise the components largely according to the approach in the Trøndelag Health Study (HUNT).^42^ ^43^ The phenotype comprises weakness, slowness, weight loss, exhaustion, and low physical activity, with detailed definitions and cut-off values provided in Supplementary Table 1. Cardiometabolic health is assessed using resting and orthostatic blood pressure, resting heart rate, and venous blood biomarkers collected at baseline and 12 months. Routine analyses include HbA1c, total and high-density lipoprotein cholesterol, creatinine and estimated glomerular filtration rate. We store additional EDTA-plasma and serum samples at −80°C in Biobank1 for potential future biomarker analyses. We assess cognitive function using the Mini-Mental State Examination (MMSE) at baseline, 6 and 12 months. During the extended follow-up, we administer the Montreal Cognitive Assessment (MoCA) at 24 and 36 months, since it additionally serves as a proxy indicator for the dementia component of disability-free survival, a prespecified exploratory outcome of the extended follow-up (Table 2). Accordingly, a single common cognitive measure cannot capture longitudinal change across the entire baseline-to-36-month period.

**Table 2.** The three components of disability-free survival.

| Endpoint | Definition and adjudication |
| --- | --- |
| Mortality | Information on mortality, including cause of death, is obtained from the Norwegian Cause of Death Registry and the Norwegian Population Registry. |
| Dementia | Dementia is ascertained from diagnoses recorded in patient records and/or through linkage to the Norwegian Patient Registry and the Norwegian Registry for Primary Health Care, where participant consent permits. For participants who have not consented to registry or patient-record linkage but complete the cognitive follow-up assessment, a Montreal Cognitive Assessment score at least 2.0 standard deviations below the age-, sex-, and education-adjusted mean is used as a proxy indicator for the dementia component of disability-free survival.<sup>54</sup> This threshold is consistent with the DSM-5 framework, in which substantial cognitive impairment in major neurocognitive disorder typically corresponds to performance at least 2 standard deviations below an appropriately adjusted normative mean, while recognising that cognitive-test performance alone does not constitute a clinical diagnosis.<sup>55</sup> |
| Physical disability | Assessed using questionnaire items covering basic and instrumental activities of daily living. Basic activities of daily living include walking indoors on the same floor, toileting, washing, bathing or showering, dressing, getting in and out of bed, and eating. Instrumental activities of daily living include preparing a hot meal, performing light and heavy housework, doing laundry, shopping, paying bills, taking medication, going outdoors, and using the bus. Physical disability is defined as being unable to perform one or more of these activities independently, without help from another person. If information on activities of daily living cannot be obtained, admission to a long-term care nursing facility is used as an indicator of physical disability. Information on long-term care is obtained from patient records and linkage to the Municipal Patient and User Register. |

Validated questionnaires assess participant-reported outcomes, including health-related quality of life, fear of falling, depressive symptoms, nutritional status, sleep quality, and musculoskeletal pain. Furthermore, habitual physical activity and muscle-strengthening exercise behaviour are assessed by self-report, while dietary intake is assessed using interviewer-administered 24-hour dietary recalls and a Norwegian translation of the Protein Screener 55+. We assess basic and instrumental activities of daily living by self-report using items adapted from established basic and instrumental activities of daily living measures, including the Physical Self-Maintenance Scale and Lawton Instrumental Activities of Daily Living Scale.^44^

For all participants, we also obtain information on healthcare utilisation, prescription and non-prescription medication use and medical outcomes from participant reports, patient records, and linkage to relevant Norwegian health registries. These data also inform health-economic analyses of the intervention.

Disability-free survival during the extended 24- and 36-month follow-up is defined as the time from randomisation to the first occurrence of death from any cause, dementia, or physical disability. Dementia is ascertained from patient records and registry linkage, or from a MoCA-based proxy indicator when linkage consent is unavailable (Table 2). Physical disability is identified from self-reported inability to perform one or more basic or instrumental activities of daily living independently or, where these data are unavailable, from admission to a long-term care nursing facility identified through patient records or linkage to the Municipal Patient and User Register (Table 2).

### Adherence

We assess adherence to the intervention protocol using attendance records and training logs. Global adherence is expressed continuously as the percentage of planned training sessions completed at the prescribed intensity and volume. A protocol-adherent session requires completing the prescribed 24 repetitions per exercise at the prescribed training intensity; from session 8 onward, the prescribed intensity is ≥80% of the most recently measured 1RM. Attendance, training intensity and training volume are also reported separately.

### Data collection methods and data management

Trained study staff collect study data using standardised procedures and study-specific case report forms. Supplementary Table 1 provides detailed procedures for the clinical assessments and outcome measures. Clinical assessment data and most patient-reported outcomes are first recorded on paper case report forms and subsequently entered into electronic datasets. We administer randomisation, monthly fall reports and quarterly questionnaires on adverse events, health, physical activity, and cognitive training through eForsk, a secure electronic data-capture platform for clinical research developed and maintained by Hemit and operated by Norsk Helsenett. According to their preference, participants either complete the questionnaires electronically or respond by telephone, with study staff entering their responses directly into eForsk. Exercise instructors document intervention delivery separately in electronic training logs.

Participants are assigned a unique study identification number, and research data are stored pseudonymised. Paper case report forms are stored in locked cabinets in locked offices, separately from signed consent forms. Electronic research data are stored in access-restricted NTNU storage solutions and eForsk, where applicable. Identifiable information, including the study identification key, is stored separately from research data and is accessible only to authorised project personnel. Data from clinical assessments, questionnaires, fall reports, training logs and registry linkages are linked using the study identification number. Registry data are obtained through applications to the relevant national registry holders and linked in accordance with applicable approvals and data protection regulations.

Data quality is supported through standardised data-collection procedures, a study-specific data dictionary, range and consistency checks, and routine review of missing or implausible values. The principal investigator or delegated study personnel oversee data-quality procedures. Study data are retained and archived in accordance with the ethics approval, institutional procedures, and applicable Norwegian regulations.

### Embedded substudies

ReFit includes embedded substudies and ancillary analyses addressing selected methodological, mechanistic, and implementation-related research questions. Substudies are conducted under separate consent procedures and are analysed and reported separately from the main trial outcomes. Participation in the main trial is not influenced by willingness to participate in substudies. All substudies must have REK approval and do not involve interventions that conflict with the main trial.

### Modifications to the protocol after trial commencement

Several protocol modifications were implemented after trial commencement. Most notably, the grip-strength eligibility criterion was revised during recruitment, first by changing the threshold from a T-score of -2.5 to -2.0 and subsequently by updating the corresponding sex-specific cut-off values using contemporary Norwegian normative data. Other modifications included allowing walking aids, adapting intervention delivery to support adherence, revising the control condition to better reflect usual care, modifying selected outcome and adverse-event assessments, and adding extended follow-up to evaluate disability-free survival and the durability of the intervention effects. The main modifications, their timing, and rationale are summarised in Table 3. Protocol modifications were submitted to REK as required and updated in the trial registry as relevant. Although some amendments affected eligibility criteria, intervention delivery, and secondary data collection, none altered the duration of the 12-month intervention or the definition and prospective ascertainment of the primary outcome: the fall rate per person-year during the 12-month follow-up.

**Table 3.** Modifications to the protocol after trial commencement.

| **Table 3. Modifications to the protocol after trial commencement** |  |  |  |
| --- | --- | --- | --- |
| **Timing** | **Area** | **Modification** | **Rationale and implications** |
| Before randomisation of the first participant | Cognitive eligibility assessment | The planned instrument for cognitive eligibility screening was changed from the Montreal Cognitive Assessment to the Mini-Mental State Examination. | Implemented before the first participant was randomised; the change affected only the cognitive screening instrument used to determine eligibility. The Montreal Cognitive Assessment was subsequently introduced as an outcome assessment when the extended 24- and 36-month follow-up was added. |
| May 2023 | Recruitment and eligibility | The eligibility criteria were revised to allow participation by individuals using walking aids, such as walkers or rollators. | Improved generalisability and recruitment feasibility among community-dwelling adults aged $\geq 80$ years. |
| December 2023 | Intervention delivery | After the first 6 months, participants allocated to the intervention group were allowed to complete one weekly unsupervised session, provided they completed at least one supervised session per week and documented their training. | Improved adherence and feasibility during the second half of the 12-month intervention period. |
| December 2023 | Control group | The control condition was modified from referral to the participant's general practitioner for follow-up to written information on current physical-activity recommendations for older adults, including fall-prevention exercises. | Referral to general practitioners was unlikely to provide standardised or trial-specific follow-up for reduced muscle strength or sarcopenia in routine care. The revised comparator improved feasibility, acceptability and standardisation while reflecting usual care, in which structured, supervised exercise programmes are typically not provided to this population. The intervention, outcomes and 12-month primary endpoint were unchanged. |
| December 2023 | Fall-related outcomes and adverse-event monitoring | Near-falls were removed from the planned outcome assessment, and the adverse-event reporting procedures were expanded to capture musculoskeletal pain and related symptoms. | Reduced participant burden and improved the clinical relevance and consistency of safety monitoring. |
| March, September 2024 | Grip-strength eligibility criterion | The grip-strength criterion for reduced muscle strength was revised during recruitment. The T-score threshold was changed from $-2.5$ to $-2.0$ relative to sex-specific normative reference values, and the | These revisions improved recruitment feasibility and the precision and relevance of the grip-strength eligibility criterion while retaining an eligibility criterion based on reduced muscle strength. Sensitivity |
|  |  | corresponding grip-strength cut-off values were later updated using contemporary normative reference data obtained with the same dynamometer model as used in the present trial. | analyses will assess the robustness of the primary findings to protocol amendments affecting eligibility criteria. |
| August 2024 | Post-trial provision for the control group | The post-trial provision for participants allocated to the control group was modified. Early in the trial, participants were offered either a gift card or 10 weeks of supervised resistance training after completing the 12-month follow-up; subsequently, most participants allocated to the control group will be offered a 1-year fitness-centre membership after completing the 12-month follow-up. | These provisions are offered only after completion of the 12-month primary outcome follow-up and are therefore not part of the primary trial comparison. They will be considered when interpreting outcomes during the extended 24- and 36-month follow-up period. |
| September 2025 | Dietary intake and exercise-behaviour assessments | Additional questionnaire-based assessments were added, including 24-hour dietary recall interviews, a Norwegian translation of the Protein Screener 55+, and the Muscle-Strengthening Exercise Questionnaire. | Enables assessment of dietary intake and muscle-strengthening exercise behaviour as potential explanatory or contextual factors. As these assessments were added during the trial, data will not be available for all participants at all time points and will be analysed and interpreted accordingly. |
| September 2025 | Extended follow-up | Follow-up assessments at 24 and 36 months were added to evaluate disability-free survival and the durability of effects on falls and selected secondary outcomes. Falls during extended follow-up will be collected using retrospective 12-month recall. | Extends follow-up beyond the 12-month primary outcome period. The intervention and prespecified 0–12-month primary endpoint were unchanged. |

### Sample size calculation

The sample size was determined *a priori* to be 240 participants (120 per group). Assuming a control-group fall rate of 1.6 falls per person-year, a mean follow-up of 0.875 person-years, an overdispersion parameter (θ) of 1.6, a two-sided α of 0.05, and analysis by negative binomial regression, this sample size gives 80% power to detect an incidence rate ratio of 0.64 – a 36% reduction in fall rate in the intervention group relative to control.

The assumed effect size was informed by a home-based strength and balance trial in older adults at high risk of falls (mean age 81.6 years), which reported a 36% reduction in falls (IRR 0.64).^45^ The overdispersion parameter (θ=1.6) was based on the value used by Liu-Ambrose et al.,^45^ derived from data reported by Shumway-Cook et al.^46^ This assumed effect size was considered clinically meaningful and is also broadly consistent with a pooled 34% reduction in the rate of falls with multiple-component exercise programmes (rate ratio 0.66).^13^ The assumed control-group fall rate lies between that of a general-population cohort aged 75+ (0.94 falls per person-year) and that of older adults selected for high fall risk after a fall (≈2.1),^45^ ^47^ reflecting the sample’s advanced age (≥80 years) and low muscle strength. The assumed mean follow-up of 0.875 person-years allows for incomplete follow-up; therefore, no additional inflation for loss to follow-up was applied. Varying observed follow-up time is accounted for using the person-time offset in the primary analysis.

### Statistical methods

This section prescribes the statistical analysis plan. We will finalise additional analytical details in a separate statistical analysis plan, to be made publicly available via ClinicalTrials.gov before database lock and before commencing outcome analyses. Primary analyses follow the intention-to-treat principle, with participants analysed according to their randomised allocation irrespective of adherence to or discontinuation of the allocated intervention.

The between-group difference in fall rate over 12 months is estimated using negative binomial regression. Treatment group, sex and age at randomisation are included as fixed effects. We report the treatment effect as an incidence rate ratio (IRR) with 95% confidence intervals.

We handle missing fall data in two ways. First, only observed data are used, with the logarithm of observed follow-up time included as an offset to account for differences in observed follow-up time. Intervals for which fall-outcome data are unavailable, and time after death or withdrawal from follow-up, do not contribute to observed person-time. Participants with no fall data at any point after randomisation remain in the intention-to-treat population but contribute no observed data to this analysis. Second, we address missing fall data using multiple imputation; the imputation model, including variables used to predict missingness and fall occurrence, will be specified in the statistical analysis plan before outcome analyses commence, and we combine estimates from the imputed datasets using Rubin’s rules.

Sensitivity analyses assess whether amendments to the eligibility criteria during recruitment influenced the estimated intervention effect. Participants are classified according to the eligibility criteria in effect at the time of enrolment. The primary model is extended to include recruitment period and a treatment group × recruitment period interaction, and treatment effects are also estimated separately within recruitment periods. We will prespecify the definition of recruitment periods and further details of this analysis in the statistical analysis plan. Additional sensitivity analyses examine the potential influence of cognitive impairment on the primary findings. We repeat the primary fall-rate analysis after excluding participants with a baseline MMSE score <24 and include baseline MMSE as a continuous covariate.

Other fall-related secondary outcomes are analysed using regression methods appropriate to their distribution: logistic regression for the occurrence of one or more falls, negative binomial regression for falls requiring medical attention and fall-related fractures, and Cox proportional hazards regression for time to first fall, each including treatment group, age at randomisation and sex as covariates.

Repeatedly measured secondary outcomes are analysed using linear or generalised linear mixed models, as appropriate for the distribution of each outcome. Group, time (treated as a categorical variable), the group × time interaction, age at randomisation, and sex are included as fixed effects, with a participant-specific random intercept included to account for repeated observations within individuals.

As a prespecified exploratory analysis, disability-free survival during the extended follow-up period is analysed using Cox proportional hazards regression, with treatment group, age at randomisation and sex included as covariates. Disability-free survival is defined as the time from randomisation to the first occurrence of death from any cause, dementia, or physical disability, as defined in Table 2. Participants without an event are censored at the end of the 36-month follow-up period. The proportional hazards assumption is assessed.

A cost-effectiveness analysis from a healthcare perspective is conducted for the 12-month intervention period. Cost-effectiveness is assessed by estimating the incremental cost-effectiveness ratio (ICER) between the intervention and control groups, with the intervention considered cost-effective if the ICER falls below the cost-effectiveness threshold.^48^ Health outcomes in the cost-effectiveness analysis are assessed using quality-adjusted life years, which combine health-related quality of life and time alive over the 12-month intervention period.^49^ Costs include intervention costs, prescription medication, and utilisation of health and care services during the 12-month intervention period. Additional details regarding the analytic perspective, valuation of costs, handling of uncertainty and missing data, and interpretation of the cost-effectiveness threshold will be specified in the health-economic analysis plan.

Additional exploratory analyses may examine associations between outcomes and attendance, adherence to the training protocol, and level of supervision. Further details of these analyses will be prespecified in the statistical analysis plan.

For the primary outcome, a two-sided p value < 0.05 is considered statistically significant. Secondary and exploratory analyses are interpreted cautiously, with model effect estimates reported in their original scale alongside 95% CIs. Where established thresholds are available, estimates are also interpreted in relation to clinically meaningful differences.

### Monitoring

Adverse events are assessed systematically every 3 months in both randomised groups using a structured questionnaire covering the preceding 3 months. Study staff administer the adverse-event questions alongside the monthly fall report at the corresponding time point, either electronically through eForsk or by telephone interview, according to participant preference; study staff enter telephone responses into eForsk. Participants report chest pain, palpitations, dyspnoea, acute infectious illness, worsening of an existing or underlying disease, pain or inflammation in joints and/or muscles, and other acute illness. Participants reporting pain or inflammation in joints and/or muscles are asked to provide additional details. Delayed-onset muscle soreness is not assessed as a separate predefined adverse-event category but may be captured within reports of musculoskeletal pain or inflammation. We document additional adverse events reported by participants or observed during exercise training or physical testing when identified. Events are recorded irrespective of their perceived relationship to study participation.

The principal investigator and/or study physician review events that raise concern for participant safety or may require medical assessment to determine whether clinical follow-up, modification, or discontinuation of study procedures is required. Serious or unexpected events considered potentially related to study procedures are reported in accordance with NTNU’s institutional incident-reporting procedures. Unreported mortality is identified through linkage to the Norwegian Population Registry. Given the intervention’s low risk and the absence of planned interim analyses, neither a data monitoring committee nor a separate steering committee is considered necessary. The investigator team provides direct trial oversight. The team monitors trial conduct informally through periodic study-team meetings, alongside the data-quality oversight described above.

Adverse events are summarised descriptively by treatment group based primarily on the quarterly adverse-event assessments, including the number and proportion of participants reporting at least one adverse event and the distribution of adverse-event categories. We report serious adverse events and events resulting in modification or discontinuation of the intervention separately. Reports from exercise training and physical testing provide supplementary information on procedure-related events. We do not plan formal between-group hypothesis testing.

## ETHICS AND DISSEMINATION

REK approved the study (reference number 462261). Separate written informed consent is obtained for participation in the extended follow-up period.

All research outputs will be communicated to participants, healthcare professionals, and the public via newsletters, social media, blog posts, meetings, and, where appropriate, plain-language summaries. Results will also be disseminated to the scientific community through publication in peer-reviewed journals and presentations at national and international conferences. Authorship of scientific publications will follow the recommendations of the International Committee of Medical Journal Editors.

De-identified participant data will not be shared due to restrictions in the ethical approval and applicable data protection regulations. Statistical code and other study materials may be made available from the corresponding author upon reasonable request.

## ANCILLARY AND POST-TRIAL CARE

After completion of the 12-month intervention period and primary outcome follow-up, participants allocated to the control group are offered a post-trial provision. Early in the trial, control participants were offered either a gift card or 10 weeks of supervised resistance training. Following discussions with user representatives, this was subsequently changed so that most control participants are offered a 1-year fitness-centre membership. The provision was intended to enhance the acceptability of control-group allocation and provide participants with an opportunity to engage in exercise after completing the primary outcome follow-up.

Because these provisions introduce post-randomisation exercise exposure in the control group, between-group comparisons at 24 and 36 months are considered exploratory. Uptake of the provision is described when interpreting extended follow-up outcomes, together with habitual physical activity and muscle-strengthening exercise behaviour (assessed at each follow-up; Supplementary Table 1). As a self-insured institution, NTNU (the trial sponsor) bears insurance liability should a participant be harmed as a result of trial-related procedures, in accordance with Norwegian law. No additional trial-related clinical care is planned beyond study assessments and adverse-event follow-up.

## DISCUSSION

ReFit addresses a critical evidence gap by testing whether high-intensity progressive resistance training, delivered as a standalone intervention, can reduce falls in the population carrying a high burden — community-dwelling older adults with reduced muscle strength. If proven effective, the findings could directly inform clinical guidelines and public health policy for a rapidly growing segment of the population, and provide the evidence base needed to implement structured resistance training programmes in community settings.

A key strength of ReFit is that it targets a population often under-represented in exercise trials despite carrying a high burden of falls, disability and healthcare use. By delivering a long-term, progressive intervention in community-based fitness centres, ReFit is designed not only to test efficacy but to generate evidence directly applicable to real-world implementation. The inclusion of cost-effectiveness analyses and extended follow-up to 24 and 36 months also allows assessment of whether any benefits are durable and economically viable — critical questions for translating trial findings into policy.

ReFit is conducted in a single geographical region in Norway, which may limit generalisability to other populations and healthcare systems. As in many exercise trials, participants and exercise instructors cannot be blinded to group allocation, and complete blinding of outcome assessors cannot be guaranteed. We define reduced muscle strength using population-specific normative reference values, resulting in relatively high absolute grip-strength thresholds. This may improve relevance to the Norwegian population but may also identify a broader group with less severe muscle weakness, potentially attenuating between-group differences. Similarly, the control group includes written physical activity guidance and fall-prevention exercises, which may further attenuate the intervention effects. Although participants with a pre-existing dementia diagnosis or an MMSE score <18 are excluded, the eligibility criteria permit inclusion of individuals with milder cognitive impairment, which may affect the accuracy of self-reported falls. For the primary outcome, prospective daily recording and monthly reporting throughout the 12-month intervention period mitigate this concern, although reporting error cannot be excluded. Prespecified sensitivity analyses examine the robustness of the primary findings by excluding participants with baseline MMSE scores <24 and by additionally adjusting for baseline MMSE as a continuous covariate. Recall bias is a greater concern for the retrospective assessment of falls before trial enrolment and during the extended 24- and 36-month follow-up; analyses of falls during extended follow-up will therefore also account for baseline cognitive function. The cognitive assessment instrument also changes from the MMSE during the 12-month intervention period to the MoCA during extended follow-up, precluding direct assessment of cognitive change on the same scale from baseline to 36 months. Protocol modifications during recruitment, including revisions to the grip-strength eligibility criterion, may introduce some heterogeneity in the trial population, and some assessments added during the trial will not be available for all participants at all time points. These limitations are addressed through sensitivity analyses and transparent reporting of protocol amendments, and are considered when interpreting the findings.

Most trials demonstrating fall-rate reductions with exercise have delivered multiple-component or balance-based programmes rather than resistance training alone. The most robust evidence derives from multicomponent interventions incorporating balance and resistance training, with resistance training generally delivered at low-to-moderate intensity such as the home-based Otago-type programme evaluated by Liu-Ambrose and colleagues,^45^ while the fall-prevention efficacy of resistance training delivered as a standalone modality remains uncertain. Progressive resistance training reliably improves muscle strength and physical function in the oldest old, but such trials have generally been powered for these outcomes rather than for falls. Recent large-scale fall-prevention trials have increasingly evaluated balance-based and digitally delivered programmes, such as the eHealth StandingTall and Safe Step interventions,^50^ ^51^ rather than high-intensity progressive resistance training in adults aged ≥80 years selected for reduced muscle strength. ReFit is therefore distinguished by its focus on this population, its use of high-intensity progressive resistance training without a balance or multimodal component, its fall-rate primary outcome, and its scale; at the time of trial initiation, it was, to our knowledge, the largest resistance-training fall-prevention trial in this age group. This design permits a direct test of whether high-intensity progressive resistance training, delivered without a dedicated balance or multimodal component, can reduce falls in older adults for whom muscle weakness is likely to be an important modifiable risk factor.

## CONCLUSIONS

ReFit is designed to provide rigorous evidence on whether high-intensity progressive resistance training reduces falls in community-dwelling adults aged ≥80 years with reduced muscle strength. The findings could directly inform clinical practice and public health policy for one of the fastest-growing and most vulnerable segments of the population.

## ACKNOWLEDGEMENTS

The authors thank all participants for their contribution. We also thank the other members of the research team, Birgitte Hyldmo and Lisbeth Røe, who contribute to the execution of the ReFit-trial. We also thank Øivind Rognmo for his role as Study Director and for departmental support of the trial. The equipment and lab facilities for physical assessments are provided by NeXt Move and the Norwegian University of Science and Technology (NTNU), and the clinical measurements are obtained and analysed at the Department of Laboratory Medicine, St Olav’s Hospital. Blood samples are stored in the Regional Biobank 1® of Central Norway.

## AUTHOR CONTRIBUTIONS

JB drafted the manuscript. JB, JSC, TF, ØOS, GMWB, BD, AS, DS, MFS, and UW conceived and contributed to the design of the study and the plan for analyses. JB is the principal investigator and coordinates the study. All authors critically revised and approved the submitted manuscript.

