## Supplemental material for "High-intensity progressive resistance training to prevent falls in community-dwelling adults aged 80 years and older with reduced muscle strength: protocol for the ReFit randomised controlled trial"

**Supplementary Table 1.** Detailed description of secondary outcome measures.

| Outcome measure | Description | Timepoint |
|---|---|---|
| **Fall-related outcomes** | Fall occurrence is assessed as the number and proportion of participants experiencing at least one fall, and the number and proportion experiencing recurrent falls (≥2 falls). Time to first fall is assessed as the time from randomisation to the first recorded fall, with participants who do not fall censored at their last date of ascertainable fall-outcome data. During the 12-month intervention period, falls are recorded prospectively using daily fall calendars with monthly reporting. During extended follow-up, falls in the preceding 12 months are assessed by retrospective recall. | M6, 12, 24, 36 |
| | Falls requiring medical attention are assessed as the number of participant-reported falls for which medical attention was sought or received. Where participant consent permits, events are additionally ascertained from patient records and linkage to the Norwegian Patient Registry using the participant's national identification number. | M6, 12, 24, 36 |
| | The number of fall-related fractures, the number of participants sustaining at least one fracture, and the number sustaining multiple fractures are identified from participant reports, and, where participant consent permits, ascertained from patient records and linkage to the Norwegian Patient Registry. Fractures are classified using applicable International Classification of Diseases codes. | M6, 12, 24, 36 |
| **Muscular strength** | Maximal dynamic muscular strength is assessed as the one-repetition maximum (1RM) on a horizontal leg press machine (M456, Technogym, Italy), following a standardised procedure. The participant first completes four repetitions at the lowest resistance setting, allowing the assessor to establish technical proficiency. The resistance is then progressively increased after each successful attempt until two unsuccessful attempts are made at the same load. A successful attempt requires both technical proficiency and a full range of motion; an unsuccessful attempt is defined as failure of either the technical or range-of-motion component. Rating of perceived exertion is recorded after each attempt and used to guide the assessor's resistance increases, such that the 1RM is typically determined within 10-15 attempts. The highest successfully lifted load is recorded as the 1RM. This standardised outcome assessment is conducted separately from 1RM testing on the Cybex Eagle leg-press machines used in the intervention. Different equipment is used for outcome assessment and training delivery for logistical reasons: the university laboratory used for outcome assessments is not equipped to deliver training sessions, and adding a separate testing day at the participating fitness centres was not feasible within the study logistics. | M0, M6, M12 |
| | Maximal isometric handgrip is assessed using a Jamar Plus+ Digital Hand Dynamometer, according to the Southampton protocol.[1] Participants sit with their backs supported, forearms resting on the chair arms, wrists in a neutral position, and thumbs facing upwards.[1] Participants alternate between the right and left hands until completing three measurements per hand. Maximal grip strength is recorded as the best of the six measurements. | M0, M6, M12, M24, M36 |
| **Muscular power** | Muscular power is assessed using a force platform (9286AA, Kistler, Switzerland) installed on the leg press machine, with loads corresponding to 60 and 80% of the leg press 1-RM. We ask participants to complete two attempts at each load (1 min rest between attempts, 2 min rest between loads) as fast and forcefully as possible during the concentric phase. Peak velocity and power are derived from force-time data for each attempt. | M0, M6, M12 |

Supplemental material

| | | |
|---|---|---|
| **Physical performance and balance** | Physical performance is assessed using the Short Physical Performance Battery (SPPB), comprising standing balance, 4-meter gait speed, and the five-times-sit-to-stand test. We record both component scores (0-4) and the total score (0-12), as well as continuous component measures.[2] | M0, M6, M12, M24, M36 |
| | Static and dynamic balance are assessed using the Mini Balance Evaluation Systems Test (Mini-BESTest).[3] For the Mini-BEST, participants complete 14 different tasks across the four domains: anticipatory postural adjustments, reactive postural control, sensory orientation, and dynamic gait. Each task is scored 0-2, with a maximum score of 28; only the worst side is used in the total score when both sides are tested.[3 4] | M0, M6, M12, M24, M36 |
| **Anthropometry and body composition** | Stature is measured without shoes using a wall-mounted Seca 222 stadiometer (Seca, Hamburg, Germany), with the mean of two measurements reported to the nearest 0.1 cm. | M0, M6, M12, M24, M36 |
| | Body mass and body composition, including fat mass, skeletal muscle mass, and appendicular skeletal muscle mass, are estimated using multi-frequency bioelectrical impedance analysis (InBody 770, Biospace, Seoul, South Korea). Participants are instructed to refrain from exercise, caffeine and nicotine use prior to testing, fast for at least four hours, and to void their bladder immediately before the body composition assessment.[5] | M0, M6, M12, M24, M36 |
| | Anthropometric measures include waist, arm, and calf circumference according to the International Society for the Advancement of Kinanthropometry Standards. The mean of two measurements reported to the nearest 0.1cm is used for analysis.[6] | M0, M6, M12, M24, M36 |
| **Cardiometabolic health** | Resting blood pressure, heart rate, and orthostatic blood pressure are assessed using an automatic blood pressure device (CasMed 740, Cas Medical Systems Inc., Branford, United States). Resting blood pressure and heart rate are measured after 5 minutes of seated rest. Three measurements are taken at 1-minute intervals on the right arm, with the mean of the last two reported. Following the final measurement, participants are asked to stand, and orthostatic blood pressure is recorded at 1 and 3 minutes post-standing. | M0, M6, M12, M24, M36 |
| | Trained personnel collect venous blood samples at baseline and 12 months. HbA1c, total cholesterol, high-density lipoprotein cholesterol and creatinine are analysed according to routine procedures at the accredited laboratory at St. Olav's University Hospital in Trondheim, Norway; estimated glomerular filtration rate is calculated from serum creatinine.<br><br>For biobanking, blood is collected in two 6 mL EDTA tubes and one 5 mL serum-separator tube with gel. Plasma and serum are processed, aliquoted and stored at −80°C in the approved general research biobank Biobank1 for potential future biomarker analyses. | M0, M12 |
| **Patient-reported outcomes** | Assessed using the Pittsburgh Sleep Quality Index (PSQI), a measure of subjective sleep quality and patterns.[7] | M0, M6, M12, M24, M36 |
| | Health-related quality of life is measured using the 12-item Short-form health survey (SF-12), which consists of 12 items across two domains: Physical Component Summary and Mental Component Summary Scores.[8] | M0, M6, M12, M24, M36 |
| | Fear of falling is assessed using the Falls Efficacy Scale-International, which measures the level of concern about falling during social and physical activities of daily living, inside and outside the home whether or not the person actually does the activity.[9] | M0, M6, M12, M24, M36 |

| | | |
|---|---|---|
| | Depression is assessed using the 30-item Geriatric Depression Scale.[10] | M0, M6, M12, M24, M36 |
| | Nutritional status is assessed using the Mini-Nutritional Assessment Short Form, a validated nutritional screening tool able to identify malnourished or at-risk-for-malnutrition older adults.[11] | M0, M6, M12, M24, M36 |
| | Musculoskeletal pain is assessed using questions adapted from the Norwegian Short Form of the Musculoskeletal Instrument for Screening Symptoms (NOSF-Miss), which cover pain intensity and interference with daily life.[12] While the full NOSF-Miss is not used, its core items are included, which reflect internationally recommended domains for clinical pain trials.[13] | M0, M6, M12, M24, M36 |
| **Frailty** | Physical frailty is assessed according to Fried's Frailty Phenotype, comprising weakness, slowness, weight loss, exhaustion, and low physical activity.[14] Components are operationalised largely according to the approach used in the Trøndelag Health Study.[15]<br><br>Weakness is defined using maximal handgrip strength, with the best of six measurements used. Weakness is defined in men as grip strength <29 kg for body mass index (BMI) ≤24 kg/m$^2$, <30 kg for BMI 24.1–26 kg/m$^2$, <30 kg for BMI 26.1–28 kg/m$^2$ and <32 kg for BMI >28 kg/m$^2$; corresponding cut-offs in women are <17 kg for BMI ≤23 kg/m$^2$, <17.3 kg for BMI 23.1–26 kg/m$^2$, <18 kg for BMI 26.1–29 kg/m$^2$ and <21 kg for BMI >29 kg/m$^2$. Slowness is defined using usual-pace 4-m gait speed, with gait speed ≤0.653 m/s in men ≤173 cm and women ≤159 cm, and ≤0.762 m/s in men >173 cm and women >159 cm classified as slowness. Weight loss is defined as self-reported weight loss >3 kg during the preceding 3 months using the Mini Nutritional Assessment–Short Form. Exhaustion is defined as a negative response to the Geriatric Depression Scale item "Do you feel full of energy?". Low physical activity is defined as reporting exercise less than once per week or never. Each component contributes one point; participants are classified as robust (0 criteria), pre-frail (1–2 criteria) or frail (3–5 criteria). | M0, M6, M12, M24, M36 |
| **Dietary intake** | Dietary intake is assessed using a combination of 24-h dietary recall interviews and a Norwegian translation of the Protein Screener 55+.[16] The 24-h dietary recall interviews are conducted through physical or telephone interviews following the same protocol as in Norkost 4.[17] | M0, M6, M12, M24, M36 |
| **Habitual physical activity level and muscle-strengthening exercise behaviour** | Habitual physical activity level is assessed using the same three questions that have been used in the Trøndelag Health Study to assess physical activity levels.[18] The three questions relate to the frequency, duration, and intensity of physical activity. For frequency, we ask participants, "How often do you exercise?", with the response options "Never", "Less than once a week", "Once a week", "2-3 times a week", or "Almost every day". For duration, we ask participants, "How long does each exercise session last?", with the response options "Less than 15 min", "15-29 min", "30 min to 1h", or "More than 1h". For intensity, we ask participants, "How hard do you push yourself?", with the response options "I take it easy, I don't get out of breath or break a sweat", "I push myself until I'm out of breath and break into a sweat", or "I practically exhaust myself". The physical activity questionnaire has previously been validated.[19] | M0, M6, M12, M24, M36 |
| | Muscle-strengthening exercise behaviour is assessed using the muscle-strengthening exercise questionnaire short form.[20] | M0, M6, M12, M24, M36 |

Supplemental material

| | | |
|---|---|---|
| **Cognition** | Cognitive function is assessed using the Mini-Mental State Examination during the intervention period and the Montreal Cognitive Assessment at follow-up assessments.[21 22] Alternate versions of the Mini-Mental State Examination are used at repeated assessments in accordance with the administration guide, to reduce potential practice effects. The Norwegian Montreal Cognitive Assessment version 8.1 is used at both extended follow-up assessments. Mini-Mental State Examination and Montreal Cognitive Assessment Scores are not treated as a single longitudinal measure of cognitive change from baseline to 36 months. | M0, M6, M12, M24, M36 |
| **Basic and instrumental activities of daily living** | Basic activities of daily living are assessed by self-report across seven activities: indoor walking, toileting, washing, bathing/showering, dressing, getting into and out of bed, and eating. Instrumental activities of daily living are assessed across nine activities: preparing a hot meal, light and heavy housework, laundry, shopping, paying bills, taking medication, going outdoors, and using public transport. Participants report whether they can perform each activity independently without assistance from another person. | M0, M6, M12, M24, M36 |
| **Healthcare utilisation** | Information on hospital admissions, outpatient clinic visits, admissions to institutional rehabilitation centres, and visits to General Practitioners (GPs), physiotherapists, and chiropractors is obtained throughout the intervention period from the Norwegian Patient Registry (NPR) and the Municipal Patient and User Register (KPR). Information on prescription medication use for the study period is obtained from the Norwegian Prescribed Drug Registry. | M0-M36 |

**Supplementary Table 2.** Exercises included in the booklet provided to the standard-care group

| Exercise | Description | Prescription |
|---|---|---|
| **Sit-to-stand** | From a seated position near the front of a chair, participants lean the trunk forwards, rise to standing in a controlled manner, and return slowly to sitting. | 10 repetitions |
| **Calf raise** | From standing, with support from a chair or bench if required, participants rise onto the toes, briefly hold the position, and lower the heels in a controlled manner. | 10 repetitions |
| **Single-leg stance** | Participants stand on one leg using as little external support as possible. | Hold 10 seconds on each leg |
| **Forward step** | From standing, participants take a long step forwards, transfer body weight onto the leading leg, and then push back to the starting position. | 10 repetitions per leg |
| **Multidirectional reaching** | From a wide standing position, participants reach upwards and laterally to one side and then bend towards the opposite side, without moving the feet. | 8-12 repetitions in each direction |
| **Tandem walking** | Participants walk forwards in a straight line with one foot placed directly in front of the other, using nearby support if needed. | 10 steps, then change direction |
| **Seated trunk rotation and stretch** | From an upright seated position, participants place one hand on the opposite knee and rotate the trunk towards the opposite side. | Hold for approximately 10 seconds; repeat twice per side |
